# Real-world effectiveness of hydroxychloroquine, azithromycin, and ivermectin among hospitalized COVID-19 patients: results of a target trial emulation using observational data from a nationwide healthcare system in Peru

**DOI:** 10.1101/2020.10.06.20208066

**Authors:** Percy Soto-Becerra, Carlos Culquichicón, Yamilee Hurtado-Roca, Roger V. Araujo-Castillo

**Author notes:** Corresponding author: Roger V. Araujo-Castillo, Av. Arenales 1302, office 310. Jesus Maria, Lima 15073, Peru., - Extension 1955.

## Abstract

**Introduction:** Peru is one of the most impacted countries due to COVID-19. Given the authorized use of hydroxychloroquine (HCQ), azithromycin (AZIT), and ivermectin (IVM), we aimed to evaluate their effectiveness alone or combined to reduce mortality among COVID-19 hospitalized patients without life-threatening illness.

**Methods:** Retrospective cohort emulating a target trial, using nationwide data of mid- and high-level hospitals from the Peruvian Social Health Insurance 01/April/2020–19/July/2020. Patients 18 yo and above with PCR-confirmed SARS-CoV-2, and no life-threatening illness at admission were included. Five treatment groups (HCQ alone, IVM alone, AZIT alone, HCQ+AZIT, and IVM+AZIT within 48 hours of admission) were compared with standard of care alone. Primary outcome was all-cause mortality rate; secondary outcomes were all-cause death and/or ICU transfer, and all-cause death and/or oxygen prescription. Analyses were adjusted using inverse probability of treatment weighting. Propensity scores were estimated using machine learning boosting models. Weighted hazard ratios (wHR) were calculated using Cox regression.

**Results:** Among 5683 patients, 200 received HCT, 203 IVM, 1600 AZIT, 692 HCQ+AZIT, 358 IVM+AZIT, and 2630 standard of care. HCQ+AZIT was associated with 84% higher all-cause death hazard compared to standard care (wHR=1.84, 95%CI 1.12-3.02). Consistently, HCQ+AZIT was also associated with higher death and/or ICU transfer (wHR=1.49, 95%CI 1.01-2.19), and death and/or oxygen prescription (wHR=1.70, 95%CI 1.07-2.69). HCQ only showed higher death and/or oxygen prescription hazard. No effect was found for AZIT or IVM+AZIT.

**Conclusions:** Our study reported no beneficial effects of hydroxychloroquine, ivermectin, azithromycin. The HCQ+AZIT treatment seems to increase risk for all-cause death.

**Funding:** Instituto de Evaluación de Tecnologías en Salud e Investigación – IETSI, EsSalud

## INTRODUCTION

The coronavirus disease 2019 (COVID-19) imposed a major global challenge given its rapid transmission worldwide and high mortality (1, 2). No pharmacological treatment have demonstrated effectiveness to control the SARS-CoV-2 infection or improving clinical outcomes, worsening the current sanitary emergency, especially for low-middle income countries (3). Traditionally, the development of any specific drug might take several years. However, given the rapid spread of the pandemic, several existing drugs have been repurposed based on in-vitro studies or low-quality evidence. Consequently, there has been extensive efforts on investigating the efficacy and effectiveness of several pharmacological treatments using randomized clinical trials as well as observational studies (4).

Currently, only dexamethasone has demonstrated benefit to reduce all-cause mortality and mechanical ventilation use; meanwhile, remdesivir seems to reduce symptoms duration and occurrence of severe adverse events (5). On the other hand, some emergently repurposed treatments have consistently demonstrated no benefits on all-cause mortality, including hydroxychloroquine with or without azithromycin, lopinavir-ritonavir, and convalescent plasma (5). Ivermectin has only demonstrated efficacy in laboratory conditions, but not evidence has yet reported on hospitalized patients with COVID-19 (6). Despite many of these studies had serious limitations, these drugs continue to being used in different health systems worldwide, and some of them have even been tested for pre-exposure prophylaxis finding the same unsuccessful results (7).

Peru has been greatly impacted by the pandemic. By October 5, 2020, it is the sixth country by number of reported cases (828 169 cases), and has the highest mortality rate worldwide (101.94 deaths per 100 000 inhabitants) (8, 9). The urgent need to address this national crisis, drove the Peruvian Ministry of Health to authorize the use of hydroxychloroquine with or without azithromycin for COVID-19 patients, later adding oral ivermectin alone or in combination with the other drugs (10, 11). Given the authorized use of these drugs, and the availability of electronic health records in the nationwide Peruvian Social Health Insurance (EsSalud), we intended to use this real-world data to emulate a randomized controlled clinical trial using propensity score weighting (12, 13). In this way, the biases traditionally associated with observational studies might be minimized producing more realistic estimates of effectiveness (14), especially when combined with novel machine-learning algorithms. Thus, our study aimed to evaluate the effectiveness of hydroxychloroquine, azithromycin, and ivermectin alone or combined, to prevent all-cause deaths in hospitalized patients with COVID-19 but without life-threatening illness. Secondarily, we evaluated two composite outcomes: death and/or intense care unit (ICU) transfer, and death and/or oxygen prescription.

## METHODS

### Study design and population

We conducted a retrospective cohort analyzing data obtained from electronic records of patients hospitalized with COVID-19 in mid- and high-level complexity hospitals from the Peruvian Social Security Health System (EsSalud). We emulated a target trial to obtain robust estimates of clinical effectiveness for hydroxychloroquine/chloroquine, azithromycin, ivermectin, alone or combined, on relevant clinical outcomes (12, 15). We retrieved anonymized data from EsSalud central data system (16) using the International Classification of Disease Tenth (ICD-10) codes for COVID-19, as defined by the Pan-American Health Organization (17).

We included patients hospitalized between April 1 and July 19, 2020 with the following criteria: 18 years old or above; confirmed SARS-CoV-2 infection by RT-qPCR; clinical manifestations compatible with non-life-threatening disease at admission (no acute respiratory failure, no sepsis or septic shock, no acute respiratory distress syndrome [ARDS], no acute pulmonary edema, no disseminated intravascular coagulation [DIC]). We excluded patients who were prescribed oxygen at admission since it is part of one of the outcomes. We also excluded patients with: self-report of pregnancy at admission; discharge, ICU admission, or death within 24 hours of admission; prescription of other experimental drugs (tocilizumab, lopinavir-ritonavir or remdesivir) within 48 hours of admission; self-reported treatment of hydroxychloroquine for rheumatological diseases; and patients with records of having received any of the studied drugs before admission.

### Treatment strategies

The standard of care was defined as antipyretics, hydration, monitorization and basic supportive care. We compared it with five treatment arms, defined as the standard of care plus: hydroxychloroquine/chloroquine alone (HCQ), ivermectin alone (IVM), azithromycin alone (AZIT), hydroxychloroquine/chloroquine plus azithromycin (HCQ+AZIT), and ivermectin plus azithromycin (IVM+AZIT) at doses recommended by the Peruvian Ministry of Health (10, 11). The decision to administer any of these treatments depended on the treating physician’s own criteria guided by the Ministry of Health recommendations, which varied over time (10, 11). Thus, there was an expected heterogeneity of regimens across different services, hospitals, and even month of hospitalization.

We allowed a grace period of 48 hours to initiate therapy to assess a more realistic clinical question: what is the effectiveness of initiating therapy compared to only receiving standard care within 48 hours of hospitalization? Hence, patients who received any of the treatment regimens after 48 hours of hospitalization were assigned to the control group, similar to an intention-to-treat analysis. As mentioned before, we only excluded patients who developed any of the outcomes within 24 hours of admission; therefore, some included patients could have developed an outcome before being assigned to any group due to the grace period (48 hours). Given that these patients could have potentially been assigned to any group, they were randomly distributed between the control and treatment groups to avoid time-dependent bias due to inappropriate exclusion or treatment assignment (12).

### Start, end of follow-up and outcomes

The onset of follow-up or time zero for each patient was the date of hospitalization. The end of the follow-up was the date of occurrence of any outcome (death, death and/or transfer to ICU, death and/or oxygen prescription), discharge, or end of follow-up by July 19, 2020. The primary outcome was all-cause in-hospital death, and the secondary composite outcomes were all-cause in-hospital death and/or transfer to ICU, and all-cause in-hospital death and/or oxygen prescription.

### Strategy for emulating random assignments

In order to emulate the random assignments of a target trial, we used a propensity score weighting for multivalued treatments employing a machine-learning approach that let us calculate balanced differences for each control and treatment groups according to their baseline covariates (18, 19). A propensity score (PS) estimates the subject probability of receiving a certain treatment given their pre-treatment characteristics, but can also be used to create weights that allows robust estimations of causal effects in observational studies (20). Moreover, we used generalized boosting models (GBM) based on machine-learning algorithms to determine which baseline covariates should be included, and to estimate the PS (15, 21). The GBM is a non-parametric approach that fits classification trees using large and non-parsimonious number of pre-treatment covariates. This improves traditional PS estimation (22) because it minimizes bias from model misspecification commonly obtained by incorrect parametric model assumptions (23), and works well with missing data (22). We trained 5000 classification trees setting the following parameters: bag fraction of 1, shrinkage factor of 0.01, test all two- and three-covariates interactions (24), and standardized mean minimization as the stop method to select optimal balance. The PS obtained were transformed into propensity score odds weights (PSW) used to estimate the average assignment effect on treated subjects (25, 26). This method sets the PSW to 1 for each treatment group participants and calculates the PSW for standard care group using the odds in the reference group (PSW = propensity score/(1-propensity score)) (26).

All the variables included in the propensity score model were selected beforehand based on expert knowledge about COVID-19, or because they were considered possible confounders, or were prognostically relevant to the outcomes (18). We did not include variables theoretically associated only with treatment assignment, but not with outcome, to avoid power reduction and/or bias amplification (18, 27). The baseline covariates were: age; sex; month of admission; healthcare center location (Capital, North, South, Centre, Rainforest); Charlson’s index at hospital admission; known comorbidities in the first 48 hours (myocardial infarct/chronic heart failure/peripheral vascular disease; chronic lung disease; mild/severe liver disease; uncomplicated/complicated diabetes mellitus; cancer, stroke/dementia/paralysis; chronic kidney disease; metabolic disease; peptic ulcer disease; HIV; uncomplicated/complicated hypertension); emergency care before admission; antibiotics (other than azithromycin) used within 48 hours of admission; previous use of angiotensin-converting enzyme inhibitors/angiotensin-II receptor antagonists; pneumonia diagnosed within 48 hours of admission. We assessed the overlap of propensity score distributions between the control group and each treatment arm to verify the common support assumption. The balance was assessed using standardized mean differences for numerical covariates, and row differences for categorical variables. We considered a threshold of 10% as indicative of meaningful imbalance (18). During balance optimization, we remained blinded to the outcome results of the study. Propensity score estimation was performed using the function *mnps* from the *Twang* package (24), and covariate balance was assessed using *cobalt* (28), both in R version 4.0.2 (29) for MS Windows Pro 10×64 bits.

### Statistical analysis

We reported baseline characteristics of the control and treatment groups and 30-days cumulative incidence estimated with the Kaplan-Meier method. Comparisons of time-to-event outcomes between treatment groups were done using weighted Kaplan-Meier (KM) survival curves. We estimated unweighted (uHR) and propensity score weighted hazard ratios (wHR) using Cox proportional hazards regression to assess effectiveness. To additionally control for residual confounding, we estimated weighted HR with doubly robust adjustment (drwHR) by performing a multivariable and weighted Cox model that incorporated all the covariates used to create the propensity score. We also performed a Bonferroni adjustment of the p-values and the 95% confidence intervals (CI) dividing the type-I-error rate of 5% by five comparisons. All survival analyses were weighted by PSWs in Stata SE version 16.1 for Windows Pro 10 x64 bits (30).

### Ethics

This study was classified as minimal risk for participants. To maintain the privacy of the patients, EsSalud’s informatics office anonymized all datasets before transfer to researchers. The protocol was approved by EsSalud’s Institutional Review Board for COVID studies (91-SGRyGIS-DIS-IETSI-ESSALUD-2020) and was also registered in the Peruvian Health Research Projects repository (PRISA, by its acronym in Spanish) with ID EI-1243 (31).

## RESULTS

We included 5683 patients from 72 healthcare centers distributed in 28 health networks at national level. Of whom 200 received HCQ, 203 IVM, 1600 AZIT, 692 HCQ+AZIT, 358 IVM+AZIT, and 2630 received standard of care (Figure 1). The 36.8% (n = 2091) of participants were women, and the age ranged from 18 to 104 years old with a mean of 59.4 years old (SD 16.3 years old). Table 1 describes the baseline characteristics of each arm of the study population.

**Table 1.**
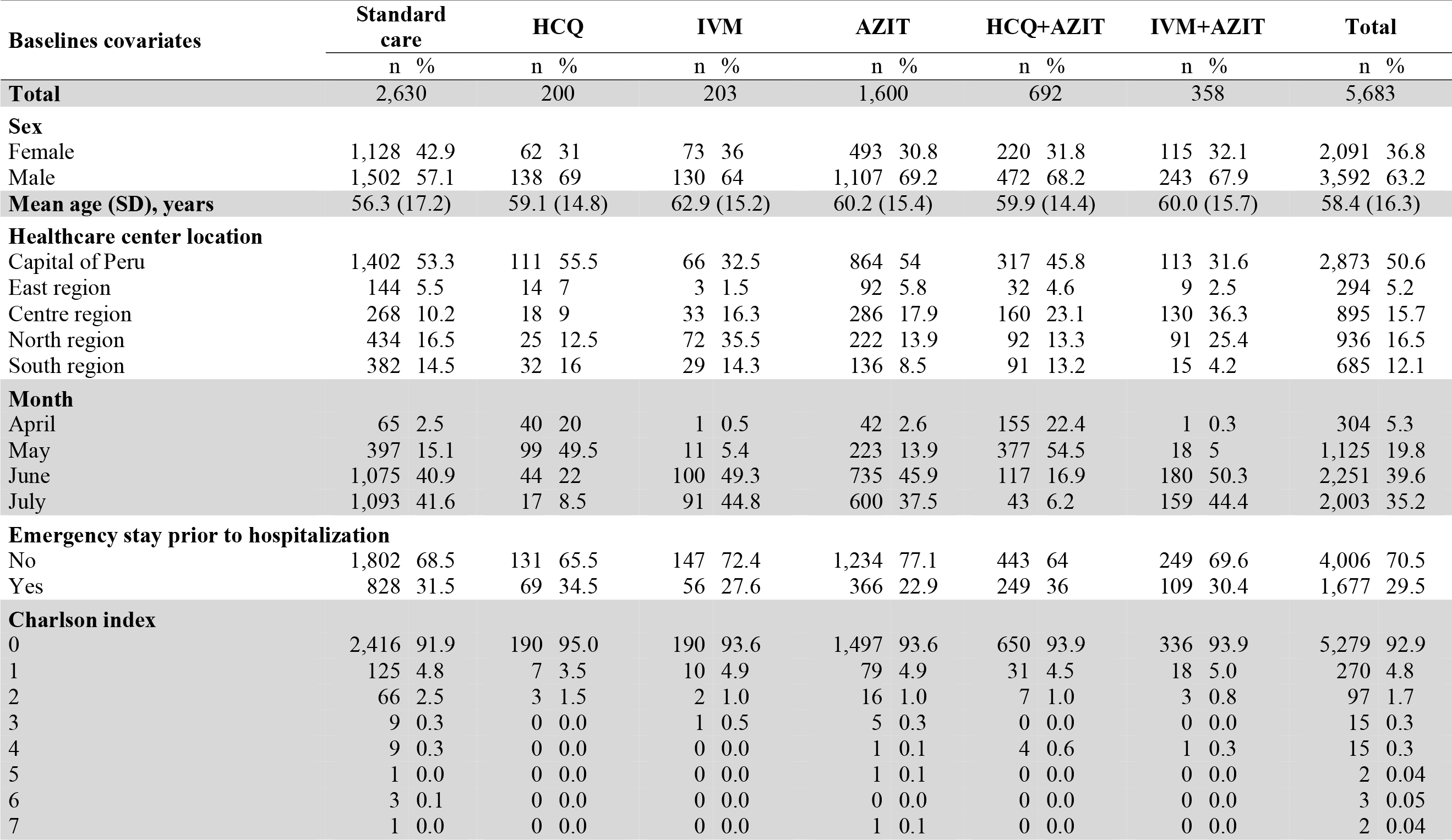
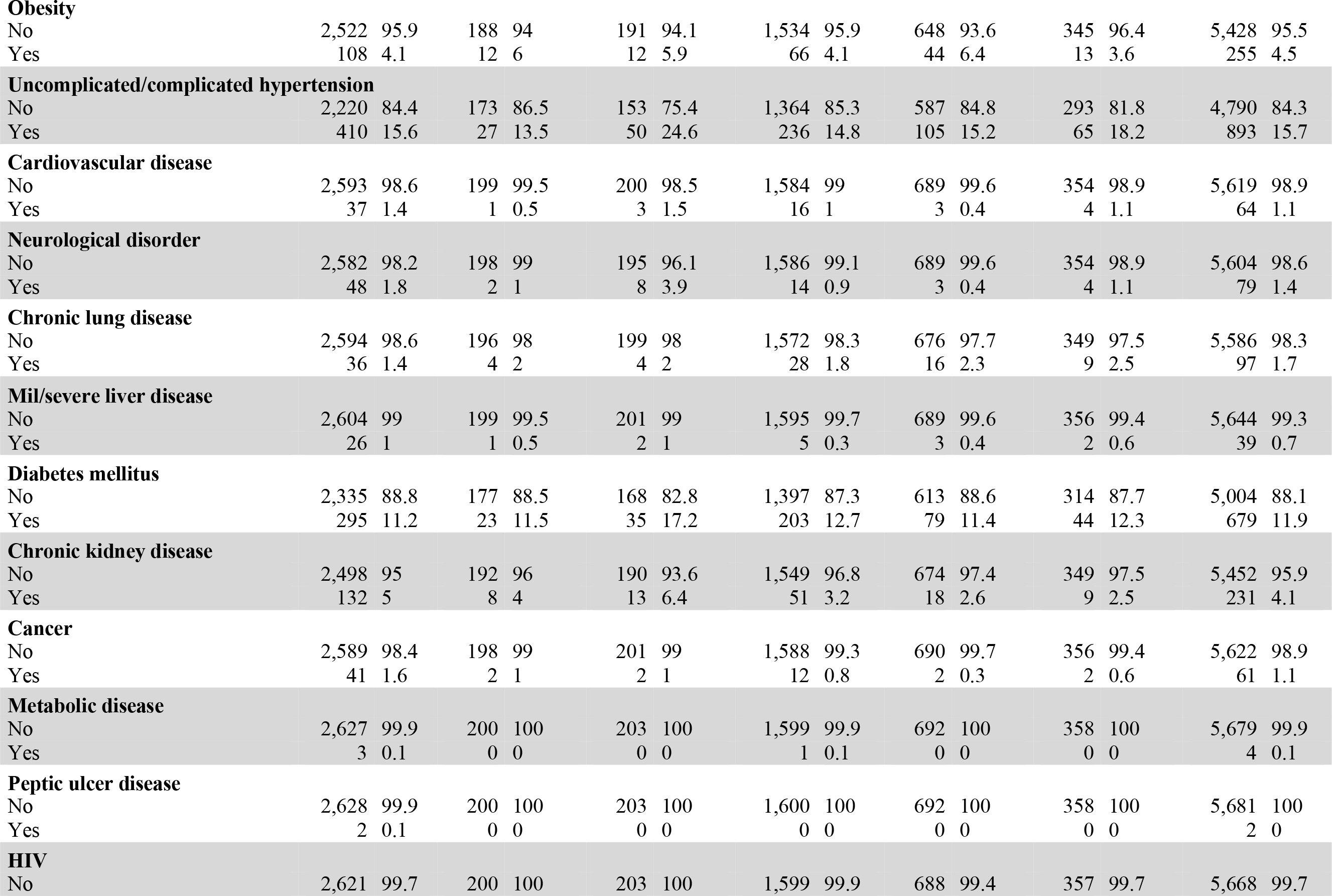
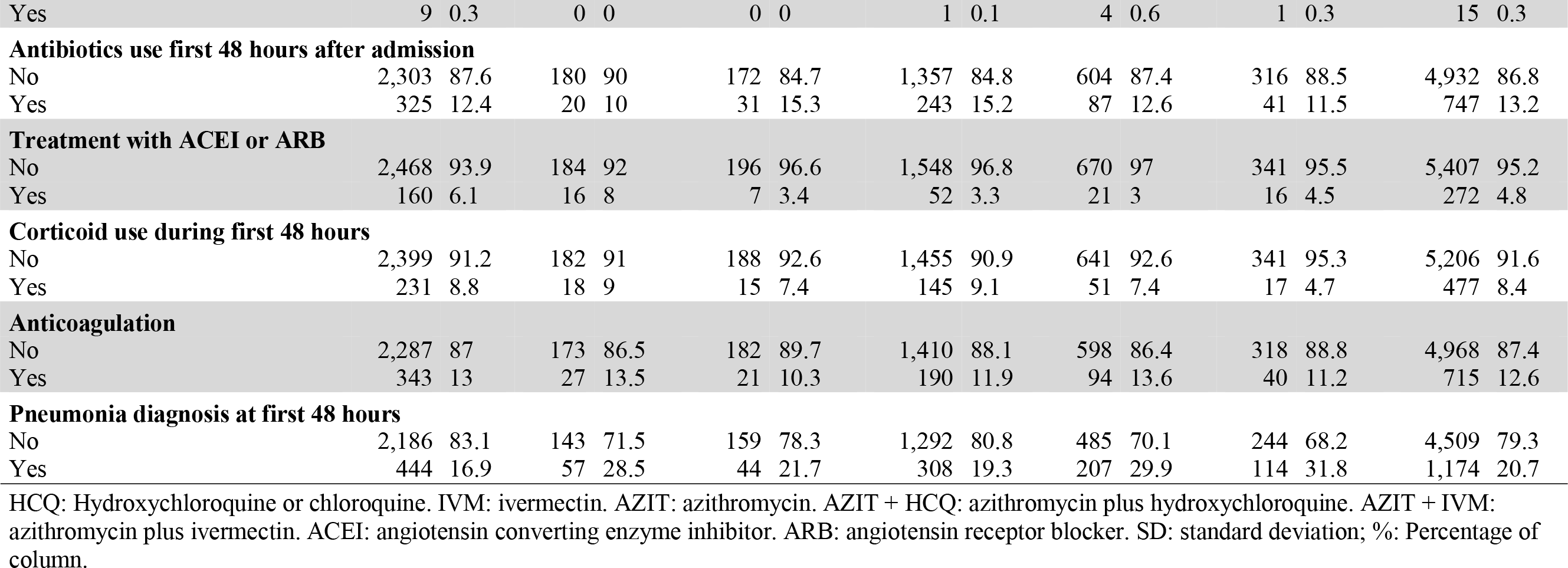
Baseline characteristics of patients with COVID-19 assigned to standard care (control group), or one of the treatment groups: hydroxychloroquine alone (HCQ), ivermectin alone (IVM), azithromycin alone (AZIT), hydroxychloroquine plus azithromycin (HCQ+AZIT), or ivermectin plus azithromycin (IVM+AZIT).

**Figure 1.**
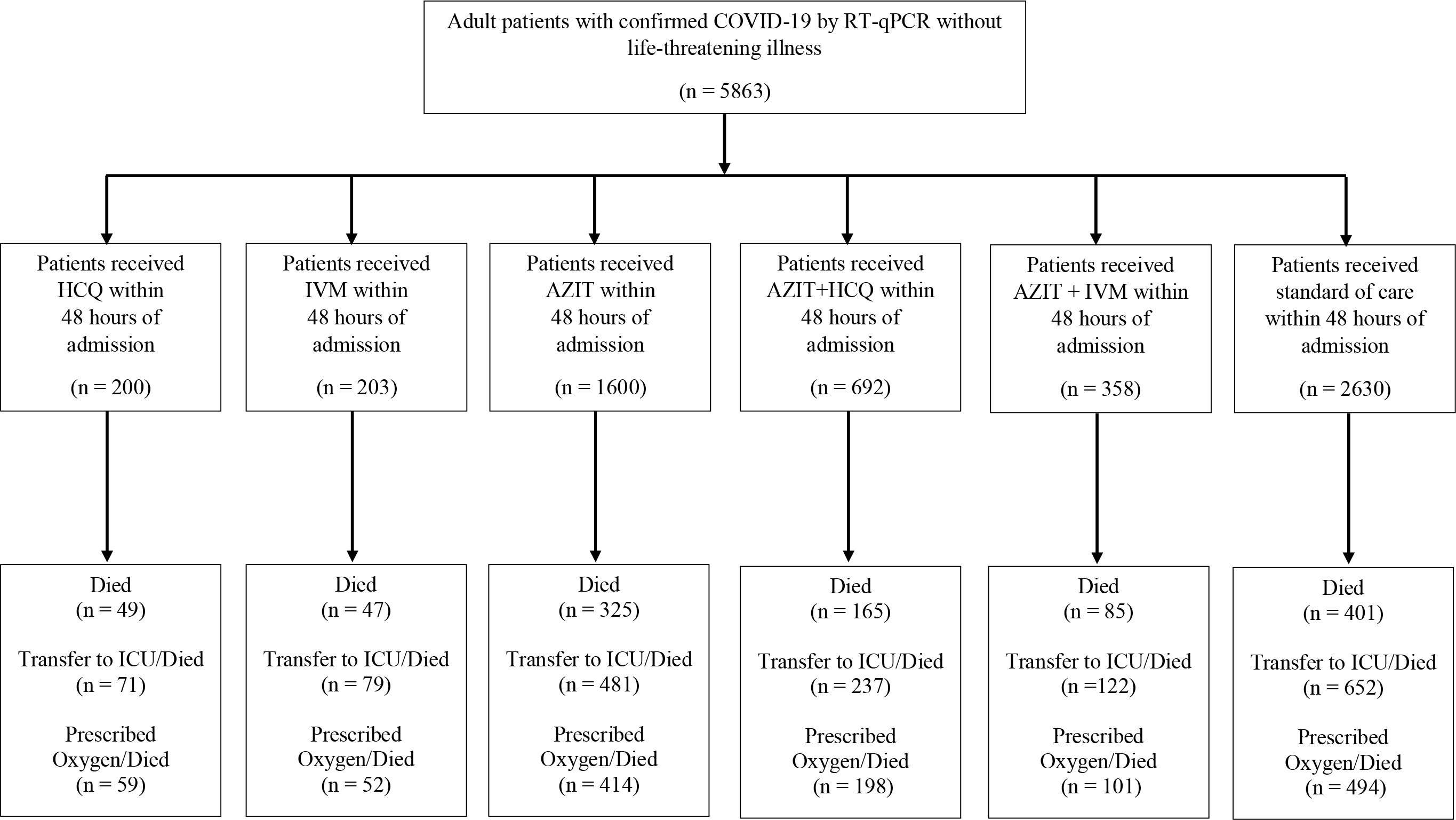
Study flowchart

### Propensity score model development

Propensity scores showed well overlapping in the region of common support between treatment groups versus control group (>95%) (SM Figure 1). Before inverse probability of treatment weighting, twelve of 30 baseline covariates had high misbalance (>10% standardized mean difference) across different groups. After applying the weighting, only one of the 30 baseline covariates (age) remained imbalanced (>10% standardized mean difference) (Figure 2).

**Figure 2.**
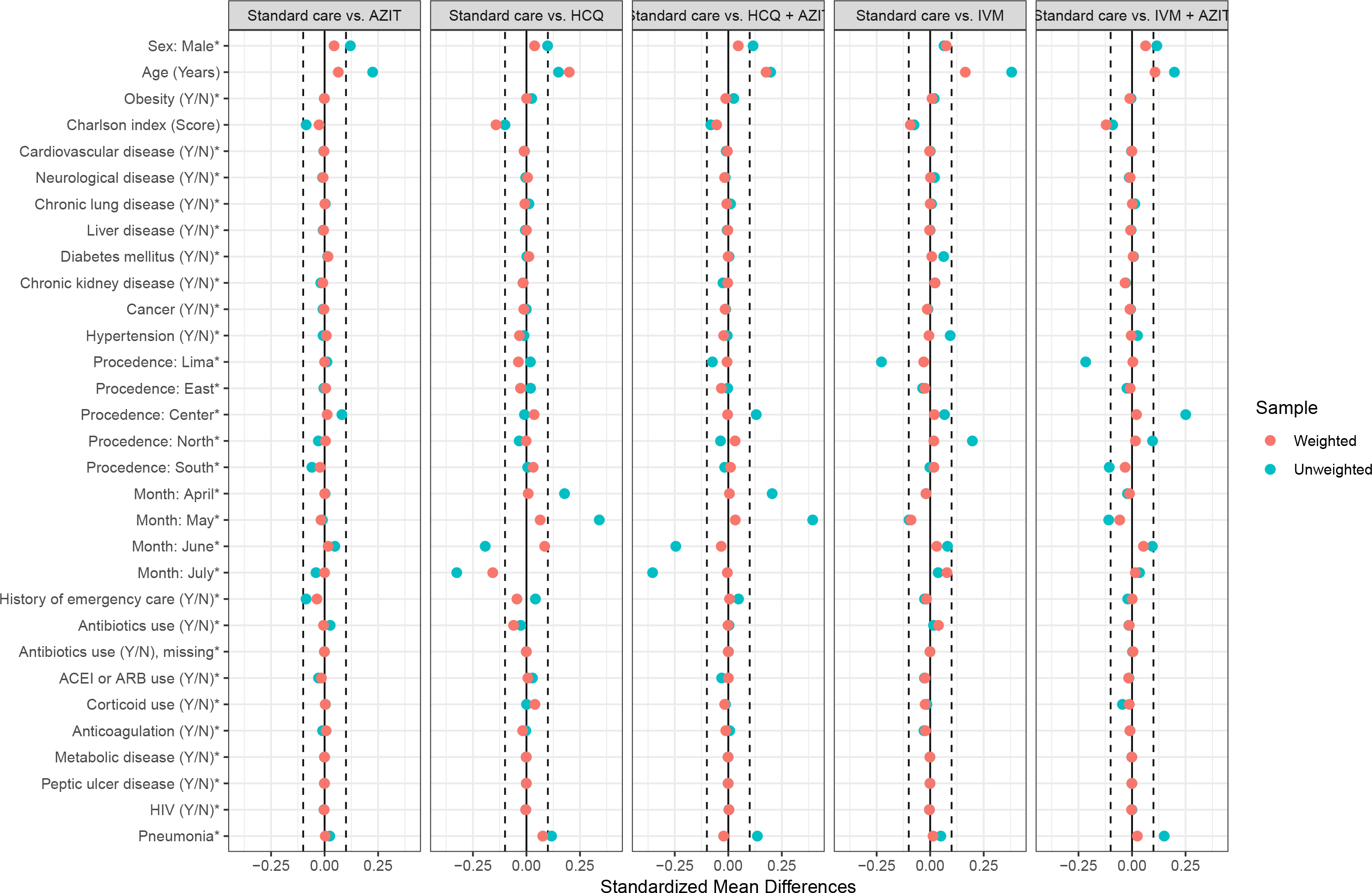
Baseline covariates balance measured as standardized mean differences, before and after inverse probability of treatment weighting

### Follow-up and outcomes

The median follow-up for overall survival was 7 days (9 for HCQ, 8 for IVM, 8 for AZIT, 9 for HCQ+AZIT, 8 for IVM+AZIT). At the end of follow-up, 1072 out of 5683 (18.9%) patients had died: 49 (15.3%) in HCQ, 47 (24.5%) in IVM, 325 (23.2%) in AZIT, 165 (23.5%) in HCQ+AZIT, and 85 (23.5%) in IVM+AZIT. Figure 3 shows weighted KM survival curves for the primary outcome, while SM Figure 2 and SM Figure 3 show weighted KM survival curves for secondary outcomes.

**Figure 3.**
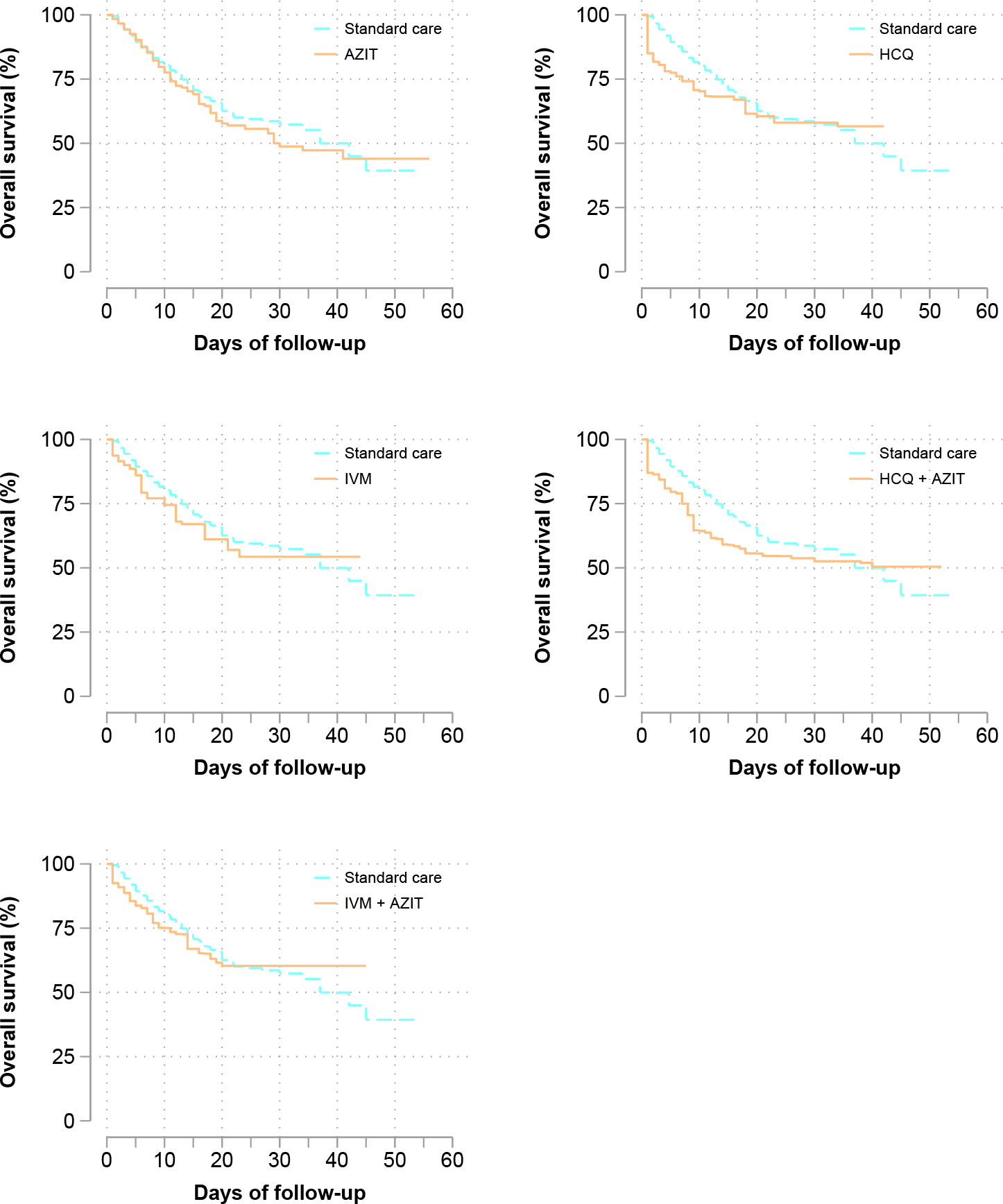
Weighted Kaplan-Meier curves for survival between each group of treatment versus standard care (control group)

Thirty-day cumulative incidence, plus unweighted and weighted hazard rates are shown in Table 2. After using the inverse probability weighting approach, we observed that HCQ+AZIT was associated with 84% higher all-cause death hazard compared to standard care (wHR = 1.84; 95%CI 1.12-3.02). Consistently, HCQ+AZIT was associated with higher all-cause death and/or ICU transfer hazard (wHR = 1.49, 95% CI: 1.01-2.19) and higher all-cause death and/or oxygen prescription hazard (wHR = 1.70, 95% CI: 1.07-2.69). Except for survival without transfer to ICU, these results were consistent even after doing double-robust adjustment to reduce residual confounding in the sensitivity analysis (see SM Table 1).

**Table 2.**
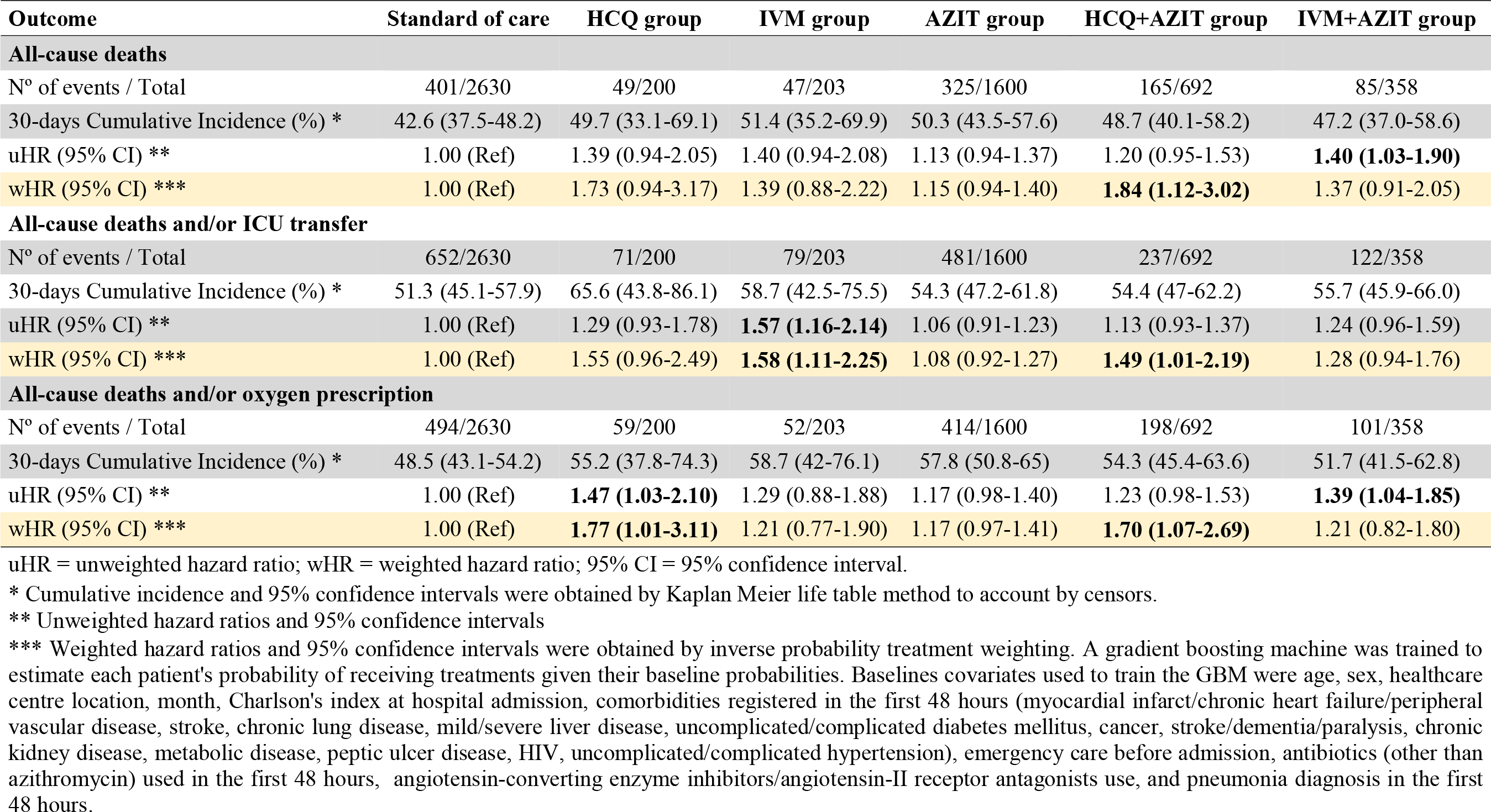
Primary and secondary outcomes in patients with COVID-19 assigned to treatment groups or standard care (control group)

We found inconsistent results regarding the estimated effect of HCQ using weighted analysis. It did not show association with the primary outcome or with death/ICU transfer, but with all-cause death and/or oxygen prescription, revealing a 77% higher hazard (wHR = 1.77, 95% CI: 1.01-3.11). The double-robust adjustment sensitivity analysis showed that HCQ treatment was associated with higher all-death hazard (drwHR = 2.08, 95% CI: 1.12-3.86), all-cause death and/or ICU transfer hazard (drwHR = 1.69, 95% CI: 1.04-2.76), and all-cause death and/or oxygen prescription hazard (drwHR = 2.13, 95% CI: 1.20-3.77) compared to the control group.

We observed that IVM was associated with higher all-cause death and/or ICU transfer hazard in the weighted analysis (wHR = 1.58, 95% CI:1.11-2.25) but not with the other two outcomes. Surprisingly, this finding persisted in the sensibility analysis (drwHR = 1.60, 95% CI: 1.12-2.27). On the other hand, we did not find effect on all-cause deaths or the composite endpoints for neither AZIT nor IVM+AZIT in all weighted (Table 2) and double-adjusted weighted analyses (SM Table 1).

## DISCUSSION

This is the first study in Latin America, a region widely impacted by the pandemic, emulating a clinical trial based on observational data comparing different drug treatments for COVID-19. Using a database based on thousands of electronic clinical records, it was possible to replicate conditions of a clinical trial under real-world conditions for several drugs prescribed during the pandemic. This study also employed novel statistical tools to emulate adequate randomization of the patients. Not only a propensity score weighting was used to balance the control and intervention groups, but they were obtained through generalized boosting models based on repeated decision trees. The resulting models for our main outcome, all-cause in-hospital deaths, and for the two secondary composite outcomes, showed no benefit from any of the treatment arms compared with standard care. There was even a consistent increase of risk developing the outcome with the hydroxychloroquine and azithromycin combination. It is then possible that the drugs evaluated have no effect on hospitalized persons who are at risk or have already developed lung involvement.

### Azithromycin alone

Our study showed no effect of azithromycin alone over mortality, survival without ICU, and survival without oxygen requirement. Few published studies have compared azithromycin alone, versus standard of care. Albani et al followed a cohort of 1403 patients either receiving azithromycin alone, hydroxychloroquine alone, the combination of both, or none of them. Using propensity score weighting, they found that azithromycin alone was associated with lower mortality (OR 0.60; 95%CI 0.42-0.85) compared to no treatment. Guerin et al (32) compared 34 patients on azithromycin alone with no treatment, finding reduction in days to achieve clinical recovery (12.9 vs 25.8; p=0.015). On the other hand, Geleris et al (33) analyzed a large single-center cohort in NYC using propensity score matching to evaluate hydroxychloroquine, but also azithromycin alone vs standard of care, finding no benefit for death and/or ICU transfer (HR 1.03; 95%CI 0.81-1.31). Arshad et al (34) also evaluated azithromycin alone as a secondary aim in their multicenter, retrospective, propensity score matched observational study, finding no effect (HR 1.05; 95%CI 0.68-1.62). Rodriguez-Molinero et al followed a cohort of 239 patients treated with azithromycin alone (35), matching 29 patients on azithromycin alone with an equal number of controls using multiple clinical and prognosis factors. They found no difference in oxygen saturation/fraction of oxygen at 48h, and a longer time to discharge in the azithromycin group. When using the unmatched whole cohort, they found no difference in any of these outcomes. In summary, the available evidence is still contradictory, but leaning on no effect.

### Hydroxychloroquine alone

We found a slight increase of risk for death and/or oxygen requirement, but not for our primary outcome (death), or death and/or ICU transfer in our weighted models. This is consistent with the systematic review published by Fiolet et al (36) including three randomized controlled trials (RCTs), one non-randomized trial, and 25 observational studies. They included 11932 patients on the hydroxychloroquine alone group, 8081 on the hydroxychloroquine/azithromycin group, and 12930 on the control group. They found no association of hydroxychloroquine alone with mortality with a pooled relative risk (RR) of 0.83 (95%CI 0.65-1.06) for all 17 studies and RR of 1.09 (95%CI 0.97-1.24) for the three RCTs. Among these studies, Geleris et al (33), after using propensity score matching, found no significant association between hydroxychloroquine and intubation or death (HR 1.04; 95%CI 0.82-1.32). On the other hand, hydroxychloroquine was associated with all three outcomes in our double-adjusted models. These results are in accord with the recently published RCT on hydroxychloroquine by the Recovery Collaborative Group (37). They found less probability of being discharged alive from hospital within 28 days (RR 0.90; 95%CI 0.83-0.98) with hydroxychloroquine, and a higher frequency of invasive mechanical ventilation or death (RR 1.14; 95%CI 1.03-1.27).

### Hydroxychloroquine/azithromycin combination

Noticeably, we found a consistent increase on the risk for the three outcomes among patients who received the hydroxychloroquine/azithromycin combination compared with standard of care. Similarly, the systematic review by Fiolet et al (36) found that hydroxychloroquine/azithromycin was associated with an increased mortality (RR 1.27; 95%CI 1.04-1.54) using six observational studies plus a RCT. This RCT (38), 447 patients, showed no improvement of clinical outcomes with the use of azithromycin in addition to standard of care that included hydroxychloroquine (OR 1.36; 95%CI 0.94-1.97) in severe COVID-19. Two other RCTs, not included in the Fiolet review, also showed no benefit. A Brazilian multicenter open-label RCT (39), 504 patients, found no difference on a seven-point ordinal scale at 15 days either with hydroxychloroquine alone, or in combination with azithromycin. More important, they found more episodes of QT prolongation and liver-enzyme elevation among patients receiving hydroxychloroquine, alone or with azithromycin. An open-label RCT in Iran compared 55 patients on hydroxychloroquine plus lopinavir with 56 patients who received azithromycin on top of that regimen, finding no difference in mortality (40). However, there are at least two big observational studies, which found a beneficial effect of the combination. Arshad et al (34) included 2561 patients, and found a 66% HR reduction with hydroxychloroquine, and a 71% reduction with hydroxychloroquine/azithromycin. Lauriola et al reported an observational study with 377 patients, finding a reduced in-hospital mortality with the hydroxychloroquine/azithromycin combination (HR 0.265; 95%CI 0.17-0.41) (41). Therefore, it is still not clear which is the real effect of the combination; however, there is a physio-pathological plausibility that the combination increased cardiac adverse events, affecting survival (42).

### Ivermectin

Until the date of this manuscript, there are no published clinical trials or large observational studies analyzing the effect of ivermectin on patients hospitalized with COVID-19. We found no association of ivermectin with all-cause mortality or with death and/or oxygen requirement; however, a deleterious effect was found on death and/or ICU admission. The reason of this association is not immediately clear for us, given the no-effect on the other two outcomes. One possibility is the presence of residual confounding despite the propensity score matching and further model adjustments. We only found pre-printed observational studies evaluating the effectiveness of this drug. The largest series is the ICON study, USA (43). They compared 173 patients on ivermectin versus 107 under usual care. They found less mortality in the ivermectin group (OR 0.52; 95%CI 0.29-0.96) and even greater effect on the subgroup with severe pulmonary disease (OR 0.15; 95%CI 0.05-0.47). A pilot study in Iraq compared 71 patients receiving hydroxychloroquine/azithromycin with 16 patients receiving ivermectin in addition (44). They found no difference on mortality (2/71 vs 0/16), but less hospitalization time in days (13.2±0.9 vs 7.6±2.8, p<0.001). Finally, there is a descriptive study in Argentina with 167 patients and no control group. They evaluated a treatment protocol that includes ivermectin, and reported an overall mortality of 0.59%, lower than their country average (45). Therefore, our study is the largest series assessing ivermectin effect among hospitalized patients and employing an adequate comparison technique.

### Limitations and Strengths

Despite being a trial emulation, this study still is an observational retrospective cohort. Without a random assignment, residual confounding is a possibility. To control this, we used a robust approach based on incident users, defining a significant time zero to prevent immortal time-bias, allowing a grace period inclusion, and using modern data science techniques to emulate random assignment. Especially, the machine learning approach (generalized boosting model) allow us to include many more potential confounders (∼30 covariates) in the weighting model, than a conventional logistic regression would allow. This is reflected in the appropriate balance and overlapping achieved between treatment and control groups once the weighting scores were applied. Moreover, a sensitivity analysis was performed using doubly robust adjustment in the weighted Cox regression models, obtaining consistently estimates of causal effects. Despite all these robust strategies, we cannot guarantee that there is some degree of residual confusion in our study. Specifically, our finding that IVM could be associated with an increased risk of one of the secondary outcomes, but not with the rest, could be due to residual confusion not properly controlled even after double adjustment.

Other possible limitation is the occurrence of non-registered variations of the treatment, as we relied on electronic records of drug dispensing. This process is strictly monitored and even audited; however, the system does not account for unexpected missing doses. On the other hand, it is important to highlight that our study, like an intention-to-treat approach, allows to estimate the effectiveness (effect in real conditions) of the studied drugs. This approach includes drug discontinuation or regimen modification due to adverse events, poor tolerability, or simply non-adherence. Thus, our results are a good approximation to the effectiveness of these treatments, but they do not necessarily reflect their efficacy (effect under ideal conditions). Although the studied drugs were administered based on the Peruvian Minister of Health guidelines, wide variations between different healthcare centers were expected. We are confident that our weighting strategy controlled most of the heterogeneity introduced by inter-hospital disparities; however, some residual confusion is still possible.

Despite these limitations, to the best of our knowledge, this is the first clinical trial emulation done in Latin America that assessed the real-world effectiveness of different treatments for COVID-19, and so far, the largest performed using robust methods to balance groups. Different from traditional observational studies, a clinical trial emulation allows for robust design, yielding reliable results. However, equilibrating control and intervention groups is no easy task. Since most common methods have disadvantages as sample size reduction (matching) or poor overlapping (weighting), the use of machine learning algorithms based on iterative decision trees (boosting) offers an excellent opportunity to optimize the balance between groups, maintaining stable models. Besides, the use of large observational data from electronic health records provides enough power to compare different treatment arms simultaneously, does not require the logistics of a RCT, and approximates treatment effect under real-world conditions.

### Conclusions

The results of this clinical trial emulation match the findings of previous RCTs and observational studies among hospitalized patients with COVID-19, which mostly showed no beneficial effects of hydroxychloroquine, ivermectin, azithromycin, or their combinations. Once assignation bias and possible confounders are controlled, the effect of the drugs studied is not significant, implying that any effect perceived in observational studies and case series is probably due to confounding effect and selection bias. We even detected a consistent increase in death risk, as well as transfer to ICU and oxygen requirement, with the hydroxychloroquine/azithromycin combination. This association has been reported in other studies and clinical trials, corroborating a potentially harmful effect of this combination. However, we did not have an adequate registry of pharmacological side effects in our electronic database, to suggest adverse reactions to this combination as a path for mortality increase.

## Data Availability

The data supporting the findings of this study are available in the IETSI EsSalud computer files,
but restrictions apply to the availability of these data, which were used under the current
study license and are therefore not publicly available. However, the data are available to the
authors upon reasonable request and with the permission of IETSI EsSalud.

## ACKNOWLEDGMENTS

We thank Engr. Pedro Vasquez, Engr. Elard Pastor and all the team from the EsSalud’s informatics office (GCTIC - Gerencia Central de Tecnologías de lnformación y Comunicaciones) for extracting and structuring the anonymized data from the electronic medical records.

## AUTHORS’ CONTRIBUTIONS

PSB conceived and designed the research idea, wrote the research protocol, conceived and designed the analysis, compiled and organized the data, performed the analysis, interpreted the data, wrote the paper, and reviewed the last version of the manuscript. Carlos Culquichicon compiled and organized the data, performed the analysis, contributed to the writing of the paper, and reviewed the last version of the manuscript. Yamilee Hurtado-Roca conceived and designed the research idea, contributed with analysis tools, interpreted the results, contributed to the writing of the paper, and reviewed the last version of the manuscript. Roger V. Araujo-Castillo conceived and designed the research idea, designed the analysis, contributed with analysis tools, interpreted the results, wrote the paper, and reviewed the last version of the manuscript.

## ETHICS STATEMENT

This study has followed all the recommendations of the Declaration of Helsinki regarding research involving human subjects. The protocol was approved by EsSalud’s Institutional Review Board for COVID studies.

