## Supplementary Material for "Real-world effectiveness of hydroxychloroquine, azithromycin, and ivermectin among hospitalized COVID-19 patients: results of a target trial emulation using observational data from a nationwide healthcare system in Peru"

**SUPPLEMENTARY MATERIAL (SM)**

SM Figure 1. Common support assessment of propensity scores in the treatment groups against standard care (control group) (n = 5683)

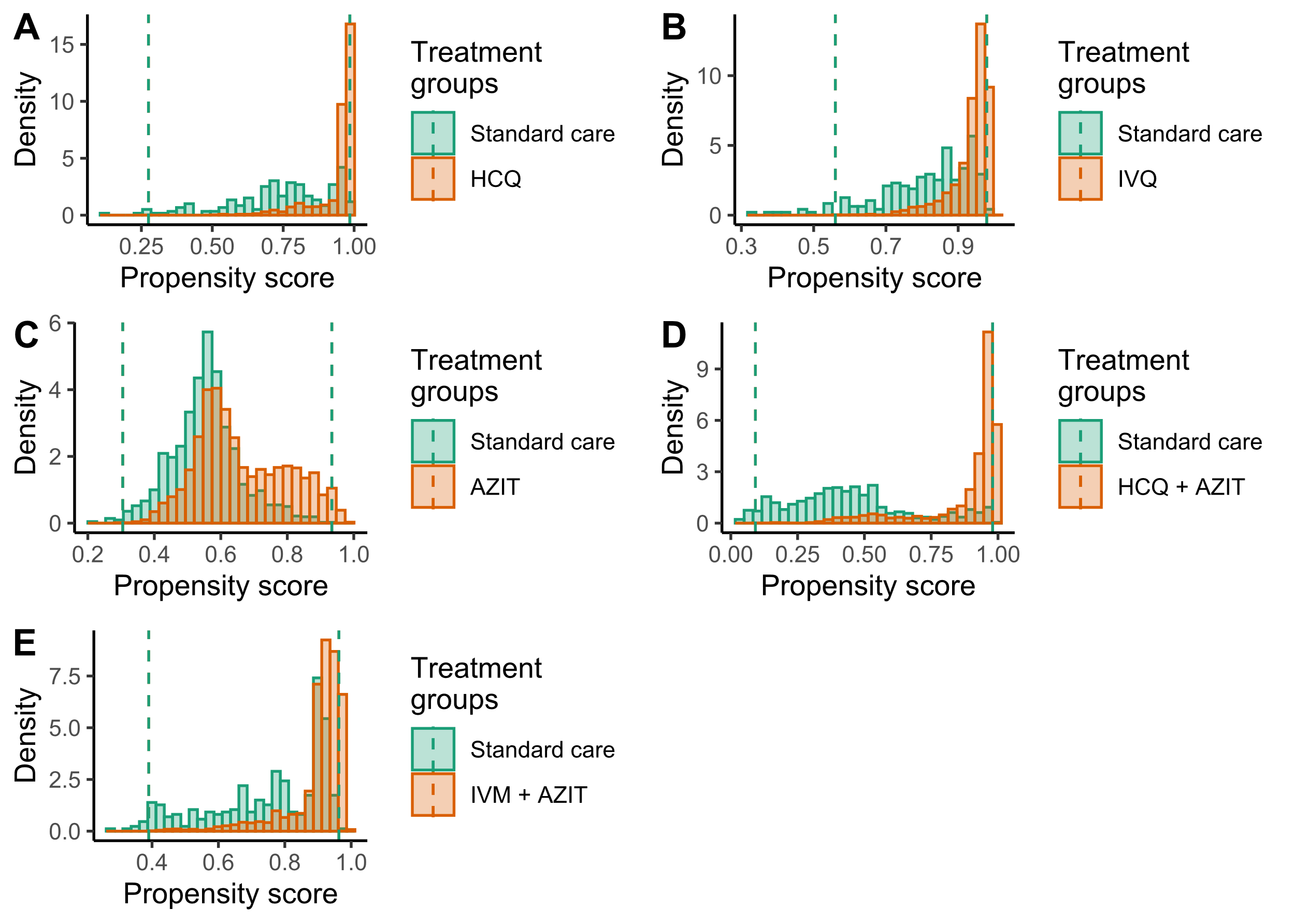

SM Figure 2. Weighted Kaplan-Meier curves for survival without ICU transfer between each group of treatment versus standard care (control group)

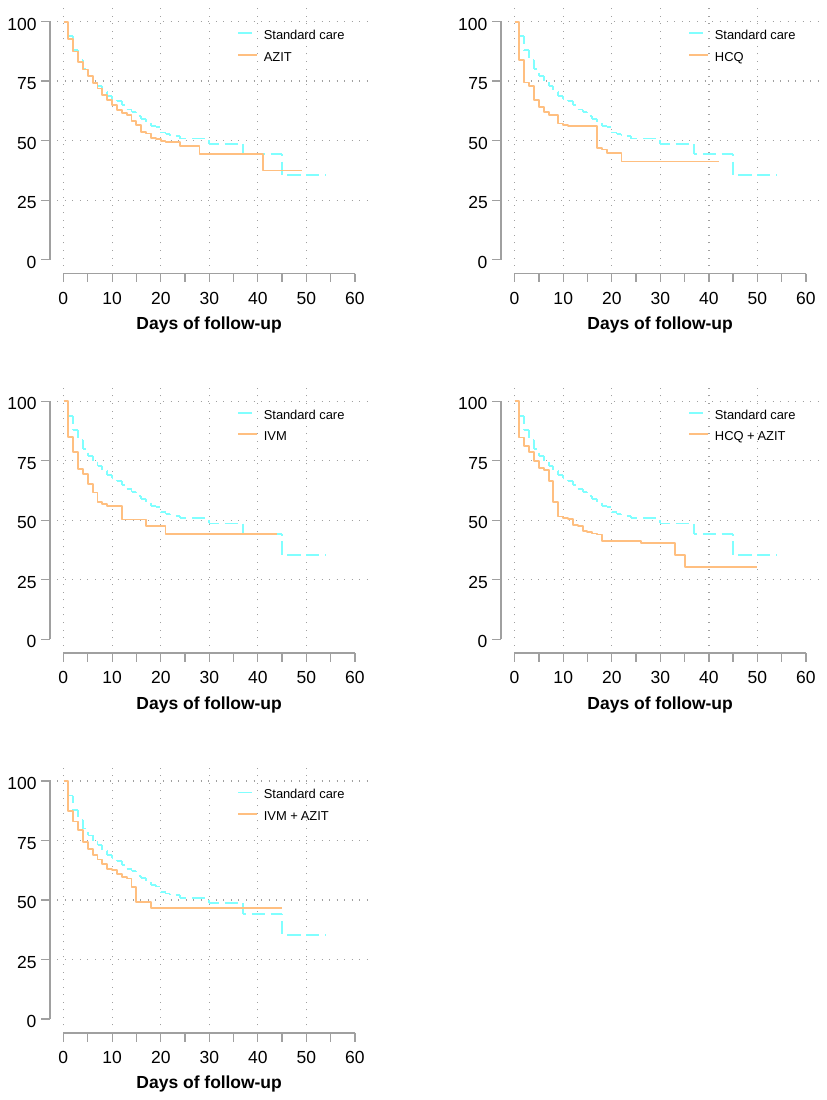

SM Figure 3. Weighted Kaplan-Meier curves for survival without oxygen prescription between each group of treatment versus standard care (control group)

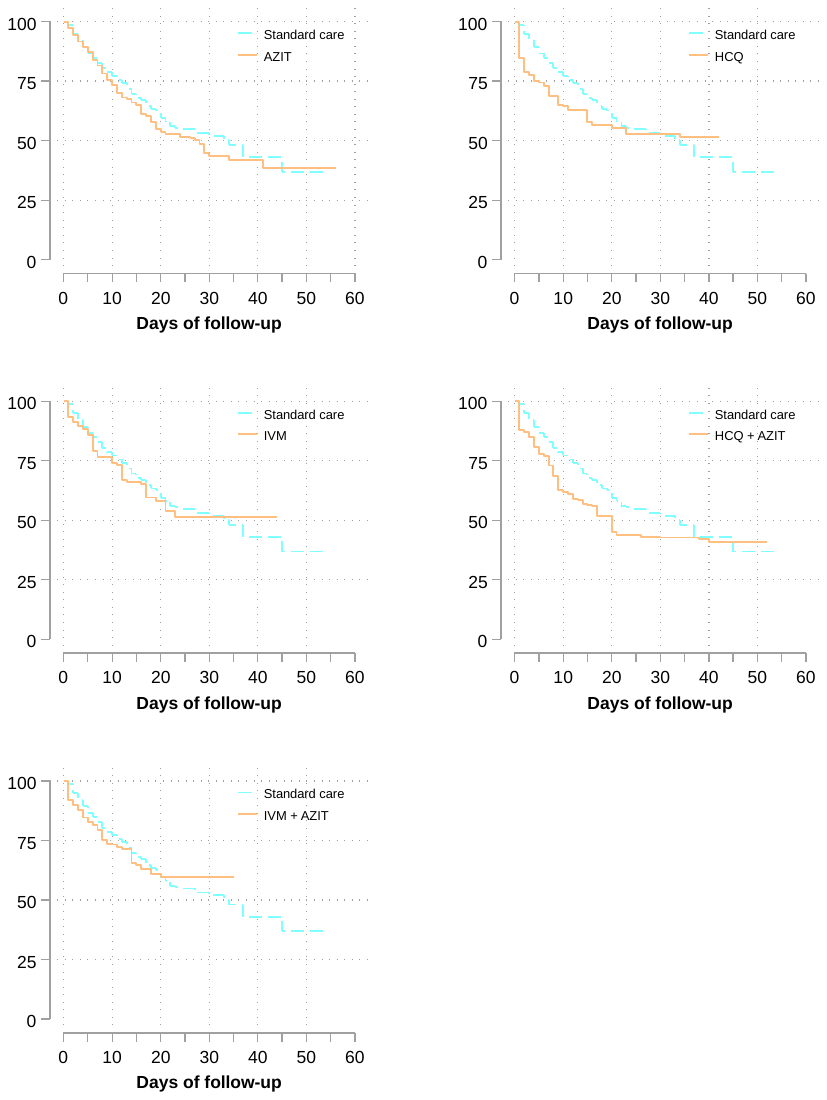

**SM Table 1. Sensitivity analysis with double robust adjusting**

| **Outcome** | **Standard care** | **HCQ group** | **IVM group** | **AZIT group** | **HCQ+AZIT group** | **IVM+AZIT group** |
| --- | --- | --- | --- | --- | --- | --- |
| **All-cause deaths** |  |  |  |  |  |  |
| Nº of events / Total | 401/2630 | 49/200 | 47/203 | 325/1600 | 165/692 | 85/358 |
| 30-days Cumulative Incidence (%) * | 42.6 (37.5-48.2) | 49.7 (33.1-69.1) | 51.4 (35.2-69.9) | 50.3 (43.5-57.6) | 48.7 (40.1-58.2) | 47.2 (37.0-58.6) |
| uHR (95% CI) ** | 1.00 (Ref) | 1.39 (0.94-2.05) | 1.40 (0.94-2.08) | 1.13 (0.94-1.37) | 1.20 (0.95-1.53) | **1.40 (1.03-1.90)** |
| wHR (95% CI) *** | 1.00 (Ref) | **2.08 (1.12-3.86)** | 1.48 (0.93-2.34) | 1.19 (0.97-1.46) | **1.83 (1.12-2.99)** | 1.54 (1.00-2.34) |
| **All-cause deaths and/or ICU transfer** | | |  |  |  |  |
| Nº of events / Total | 652/2630 | 71/200 | 79/203 | 481/1600 | 237/692 | 122/358 |
| 30-days Cumulative Incidence (%) * | 51.3 (45.1-57.9) | 65.6 (43.8-86.1) | 58.7 (42.5-75.5) | 54.3 (47.2-61.8) | 54.4 (47-62.2) | 55.7 (45.9-66.0) |
| uHR (95% CI) ** | 1.00 (Ref) | 1.29 (0.93-1.78) | **1.57 (1.16-2.14)** | 1.06 (0.91-1.23) | 1.13 (0.93-1.37) | 1.24 (0.96-1.59) |
| wHR (95% CI) *** | 1.00 (Ref) | **1.69 (1.04-2.76)** | **1.60 (1.12-2.27)** | 1.08 (0.92-1.27) | 1.37 (0.93-2.01) | 1.34 (0.97-1.85) |
| **All-cause deaths and/or oxygen prescription** | | |  |  |  |  |
| Nº of events / Total | 494/2630 | 59/200 | 52/203 | 414/1600 | 198/692 | 101/358 |
| 30-days Cumulative Incidence (%) * | 48.5 (43.1-54.2) | 55.2 (37.8-74.3) | 58.7 (42-76.1) | 57.8 (50.8-65) | 54.3 (45.4-63.6) | 51.7 (41.5-62.8) |
| uHR (95% CI) ** | 1.00 (Ref) | **1.47 (1.03-2.10)** | 1.29 (0.88-1.88) | 1.17 (0.98-1.40) | 1.23 (0.98-1.53) | **1.39 (1.04-1.85)** |
| wHR (95% CI) *** | 1.00 (Ref) | **2.13 (1.20-3.77)** | 1.26 (0.81-1.96) | 1.20 (0.99-1.45) | **1.69 (1.06-2.72)** | 1.34 (0.89-2.01) |
| uHR = unweighted hazard ratio; wHR = weighted hazard ratio; 95% CI = 95% confidence interval. | | | | | |  |
| * Cumulative incidence and 95% confidence intervals were obtained by Kaplan Meier life table method to account by censors.  ** Unweighted hazard ratios and 95% confidence intervals | | | | | | |
| *** Weighted hazard ratios and 95% confidence intervals were obtained by inverse probability treatment weighting. A gradient boosting machine was trained to estimate each patient's probability of receiving treatments given their baseline probabilities. Baselines covariates used to train the GBM were age, sex, healthcare centre location, month, Charlson's index at hospital admission, comorbidities registered in the first 48 hours (myocardial infarct/chronic heart failure/peripheral vascular disease, stroke, chronic lung disease, mild/severe liver disease, uncomplicated/complicated diabetes mellitus, cancer, stroke/dementia/paralysis, chronic kidney disease, metabolic disease, peptic ulcer disease, HIV, uncomplicated/complicated hypertension), emergency care before admission, antibiotics (other than azithromycin) used in the first 48 hours, angiotensin-converting enzyme inhibitors/angiotensin-II receptor antagonists use, and pneumonia diagnosis in the first 48 hours. | | | | | | |
